# Reconfiguration of Amplitude Driven Dominant Coupling Modes (DoCM) mediated by α-band in Adolescents with Schizophrenia Spectrum Disorders

**DOI:** 10.1101/2020.06.02.20118851

**Authors:** Stavros I. Dimitriadis

**Affiliations:** Cardiff University Brain Research Imaging Centre, School of Psychology, Cardiff University, Cardiff, United Kingdom; Neuroinformatics Group, Cardiff University Brain Research Imaging Centre, School of Psychology, Cardiff University, Cardiff, United Kingdom; Division of Psychological Medicine and Clinical Neurosciences, School of Medicine, Cardiff University, Cardiff, United Kingdom; School of Psychology, Cardiff University, Cardiff, United Kingdom; Neuroscience and Mental Health Research Institute, School of Medicine, Cardiff University, Cardiff, United Kingdom; MRC Centre for Neuropsychiatric Genetics and Genomics, School of Medicine, Cardiff University, Cardiff, United Kingdom

**Keywords:** EEG, multiplexity, chronnectomics, schizophrenia spectrum disorders, cross-frequency coupling

## Abstract

Electroencephalography (EEG) based biomarkers have been shown to correlated with the presence of psychotic disorders. Increased delta and decreased alpha power in psychosis indicate an abnormal arousal state. We investigated brain activity across the basic EEG frequencies and also dynamic functional connectivity of both intra and cross-frequency coupling that could reveal a neurophysiological biomarker linked to an aberrant modulating role of alpha frequency in adolescents with schizophrenia spectrum disorders (SSDs).

A dynamic functional connectivity graph (DFCG) has been estimated using the imaginary part of phase lag value (iPLV) and correlation of the envelope (corrEnv). We analyzed DFCG profiles of electroencephalographic resting state (eyes closed) recordings of healthy controls (HC) (n=39) and SSDs subjects (n=45) in basic frequency bands {δ,θ,α_1_,α_2_,β_1_,β_2_,γ}. In our analysis, we incorporated both intra and cross-frequency coupling modes. Adopting our recent Dominant Coupling Mode (DoCM) model leads to the construction of an integrated DFCG (iDFCG) that encapsulates the functional strength and the DoCM of every pair of brain areas.

We revealed significantly higher ratios of delta/alpha1,2 power spectrum in SSDs subjects versus HC. The probability distribution (PD) of amplitude driven DoCM mediated by alpha frequency differentiated SSDs from HC with absolute accuracy (100%). The network Flexibility Index (FI) was significantly lower for subjects with SSDs compared to the HC group.

Our analysis supports a central role of alpha frequency alterations in the neurophysiological mechanisms of SSDs. Currents findings open up new diagnostic pathways to clinical detection of SSDs and supports the design of rational neurofeedback training.

**Highlights:**

- Ratios of delta/alpha_1,2_ relative power spectrum were significant higher in SSDs subjects compared to HC
- Probability distribution (PD) of amplitude driven DoCM mediated by alpha frequency differentiated SSDs from HC with 100%
- Network Flexibility index (FI) was significant lower for subjects with SSDs compared to HC group.

## 1. Introduction

Psychosis is characterized by hallucinations and delusions or other positive symptoms which have been seen as the prevalent clinical markers of psychotic disorders like schizophrenia. However, psychotic symptomatology can be seen in both general and clinical populations. Recent research has considered psychosis as a continuum process rather than a categorical ‘on/off clinical state. This dimensional clinical status of psychosis includes psychotic-like experiences, schizotypal symptoms, risk at mental states etc (Van Os et al., 2000; Yung et al., 2003). A recent meta-analytic plan suggests that the predominance of psychotic symptoms in the general population lies at 7% (Linscott and van Os, 2013). These experiences are transitory in 80% of cases, in 20% they are persistent and in 7% they predate the onset of a psychotic disorder (Kaymaz et al., 2012; Linscott and van Os, 2013; Zammit et al., 2013).

Psychotic symptoms within the schizophrenia spectrum comprise of childhood-onset schizophrenia with age of onset ≤ 12 years like schizoaffective, schizotypal and schizophreniform disorder. It is more than evident that psychotic symptoms are differentiated between children and adolescents in terms of repetition, intensity and these includes anxiety, obsessions etc (Courvoisie et al., 2001). Schizophrenia Spectrum Disorders (SSDs) and subclinical symptoms on the psychosis continuum share cognitive demographic characteristics, cognitive levels and etiological risk factors (van Os et al., 2009; Linscott and van Os, 2013). The aforementioned findings supported that psychosis is a phenotype with a persistence symptomatology and an increased severity correlated with clinical symptomatology (van Os and Linscott, 2012). From the current literature, there are no consistent findings to characterize the phenotype of sub-clinical psychosis (Kapil and Barrantes-Vidal, 2015; Rössler et al., 2015). The majority of clinical research over sub-clinical psychosis focused mainly on SSDs probably because of their consistency in the clinical symptomatology of psychosis onset (Tandon et al., 2012). It is evident that various types of experiences have not been differentiated on SSDs (Yung et al., 2009). Further, psychotic-like experiences (PLEs) which include hallunications and delusions are quantitatively similar to clinical psychotic symptoms (DeRosse and Karlsgodt, 2015). However, only a few studies attempted to explore whether the psychotic symptoms like auditory hallucinations of non-clinical populations are or not phenotypically similar to clinical populations (Hill et al., 2012).

Cortical dis-/dysconnectivity theory of schizophrenia (Friston et al., 2016) has also been explored with electrophysiological methods. The analysis of multichannel electro (EEG) and magneto-encephalography (MEG) signals provides unique information for functional interaction between neural oscillations characterizing brain activity of specific areas within the same frequency (intra-frequency coupling) and between frequencies (cross-frequency coupling; Dimitriadis et al., 2017b,2018c) at high temporal resolution (Canolty and Knight, 2010; von Stein and Sarnthein, 2000; Lisman and Buzsaki, 2008; Dimitriadis et al., 2017b, 2018c). These functional interactions are altered in schizophrenia (Allen et al., 2011). The analysis of EEG at rest demonstrated an increased connectivity pattern in δ (1 – 4 Hz), 0 (4 –8 Hz) and β (13 – 30 Hz) frequencies complementary to a decreased pattern in α frequency within the frontal cortex (“*hypo-frontality*”) in schizophrenic patients relative to controls (Ingvar and Franzen, 1974; Ragland et al., 2007). An additional potential biomarker for schizophrenia is *abnormal brain asymmetry* (Oertel-Knoechel et al., 2012; Gotts et al., 2013; Ribolsi et al., 2014; Miyata et al., 2012).

A recent EEG study at resting-state explored aberrant static functional brain connectivity induced by schizophrenia adopting three connectivity estimators, alternative network metrics and two reference systems (Olejarczyk and Jernajczyk, 2017). The whole connectivity analysis focused on five frequencies: 2–4 Hz (δ), 4.5–7.5 Hz (θ), 8–12.5 Hz (α), 13–30 Hz (β), 30–45 Hz (γ). The authors revealed the inter-hemispheric asymmetric group-difference using Directed Transfer Function granger causality estimator at resting-state. Another study proposed a methodology of selecting the best set of EEG sensors based on their connectivity profile with the rest of EEG sensors in order to design an optimal classifier for the discrimination of healthy controls (twenty five) from schizophrenic patients (twenty five) (Dvey-Aharon et al., 2017). These authors conducted connectivity analysis based on correlation between pair-wise time series in a broadband frequency of 0.1 Hz-30 Hz. They reported a classification performance of 93.8%. The main disadvantage of both studies was the use of a static connectivity analysis approach versus a more dynamic one that has the advantage of harnessing the high temporal resolution of the EEG modality. Moreover, only the first study explored static connectivity analysis within specific frequencies while both did not explore any cross-frequency interactions, missing important parts of the rich information contained in the human EEG/ MEG (Dimitriadis et al., 2015a, 2016a,b, 2018a, c,d).

A recent study revealed distinct electroencephalographic patterns of delta/alpha brain activity in psychotic disorders including schizophrenia, bipolar disorder and methamphetamine-induced psychotic disorder (Howells et al., 2018). Delta synchronisation is expressed with an increment of EEG delta activity, has been reported in ScZ and bipolar disorder (Howells et al., 2018). Patients with schizophrenia have shown reduced delta wave activity also in sleep stages 3 and 4 (Sekimoto et al., 2010) and also during the perception of neutral and emotionally salient words (Alfimova and Uvarova, 2008). Previous studies showed higher delta synchrony during resting eyes closed condition for ScZ when compared with controls (Borisov et al., 2005). Few studies in ScZ have reported relative delta synchronization during rest with open eyes or during the completion of a cognitive task (Basar et al., 2013a,b). We expect to see also delta synchronisation (increased relative power) in SSDs subjects that will be studied here (hypothesis I).

Alpha desynchronization is expressed via a decrement of alpha activity, has been reported in ScZ and bipolar disorder (Howells et al., 2018). In ScZ, alpha desynchronization is reported in un-medicated, medicated ScZ adolescent onset ScZ and first-episode ScZ and (John et al., 2002). Alpha synchronisation is a marker of healthy resting wakefulness which reflects the readiness of the brain to process salient information (Klimesch et al., 1999). The exaggerated desynchronization of alpha activity at resting-state in psychotic disorders represents probably an inappropriate readiness to attend and process salient or not information, whether internal or external (Klimesch et al., 1999). We expect to see also alpha desynchronization (decreased relative power) in SSDs subjects that will be explored here (hypothesis II). Motivated by a recent study (Howells et al., 2018), we will estimate also the ratio of Relative Signal Power (RSP) between δ frequency and the two α subbands.

A consistent decreased functional connectivity pattern in in the α-frequency band has been reported in ScZ (Di Lorenzo, et al., 2015; Tauscher et al.,1998). Interestingly, two studies reported a high correlation between functional connectivity at rest in the α-frequency band with symptoms of ScZ (Hinkley et al., 2011; Merrin and Floyd, 1996). Preliminary evidence suggests that β-band functional connectivity is influenced by illness progression and clinical symptomatology (Di Lorenzo, et al., 2015). However, there is no study that analysed also cross-frequency coupling in conjunction with within frequencies coupling using EEG in SSD.

We hypothesized that the construction of an integrated Dynamic Functional Connectivity Graph (iDFCG) with the incorporation of dominant intrinsic coupling modes (DoCM) which can be either intra or inter-frequency coupling estimations (cross-frequency couplings) will reveal significant features related to SSDs (Dimitriadis et al., 2017b, 2018c). iDFCG tabulates both the strength and the type of interaction between every pair of sensors at every temporal segment. Our analysis will focus on phase-to-phase/amplitude-to-amplitude within frequency couplings and phase-to-amplitude/amplitude-to-amplitude cross-frequency couplings. We expected that the probability distribution (PD) of DoCM related to cross-frequency interactions will be higher for SSDs patients than their healthy controls counterparts mostly in lower frequencies (Sharp and Hendren, 2007) (hypothesis III). The ratio of cross-frequency interactions versus the total number of exist functional interactions can be seen as a nonlinearity index of dynamic functional information flow indicator of resting-state. We also assumed that SSDs patients will demonstrate a less flexible global brain pattern based on the fluctuation of DoCM across experimental time compared to healthy controls (hypothesis IV). Our final hypothesis is that amplitude and phase-based DoCM will dissociate their distinct role in both groups (hypothesis V). Our hypothesis is supported by recent evidences about the sensitivity of phase-based intrinsic coupling modes in disorders with functional alterations while amplitude-based intrinsic coupling modes showed predominant sensitivity to structural alteration (Engel et al., 2013). Here, we attempted to detect consistent patterns of brain activity and connectivity in adolescents with SSDs with a non-invasive method, EEG, following for the very first time a multiplex scenario via our DoCM model. Our analysis will dissociate the role of amplitude and phase based connectivity patterns under this framework in SSDs.

## 2. Material and Methods

The subjects were adolescents who had been screened by psychiatrist and divided into a healthy group (*n* = 39) and a group with symptoms of schizophrenia (*n* = 45). Both groups included only two groups of Russian (Moscow) school children boys aged 10-14 year. The age of the group with schizophrenia spectrum disorders (SSDs) (schizophrenia (childhood-onset), schizotypic disorder, or schizoaffective psychosis) with comparatively homogeneous symptoms ranged from 10 years and 8 months to 14 years (12.3 ±1.2 years). Clinical evaluation of these adolescents with disorders of schizophrenic spectrum was provided by experts from the National Center of Mental Health of the Russian Academy of Medical Sciences. They were diagnosed according to ICD–10 in Mental Health Research Center, Moscow and it was originally consisted of 125 boys 8–15 years old. The diagnosis was schizophrenia, childish type (F20), schizotypal disorder (F21) and schizoaffective disorder (F25). For further reading, see http://protein.bio.msu.ru/~akula/korsak/Korsak-eng.htm.

The healthy control group included 39 healthy schoolboys aged from 11 years to 13 years and 9 months. The mean age of healthy control group was (12.3±1.3 years). SSD group is recorded even before the pharmacological treatment appointments while subjects have been selected with the same severity. The two groups didn’t differ on their age.

EEG activity was recorded from 16 EEG channels at resting-state with eyes closed. The electrode positions are demonstrated in Figure 1. The 16 EEG electrodes were placed according to the international 10–20 system rules at the following locations:O1, O2, P3, P4, Pz, T5, T6, C3, C4, Cz, T3, T4, F3,F4, F7, and F8 and monopolarly referenced to coupled ear electrodes.

**Figure 1.**
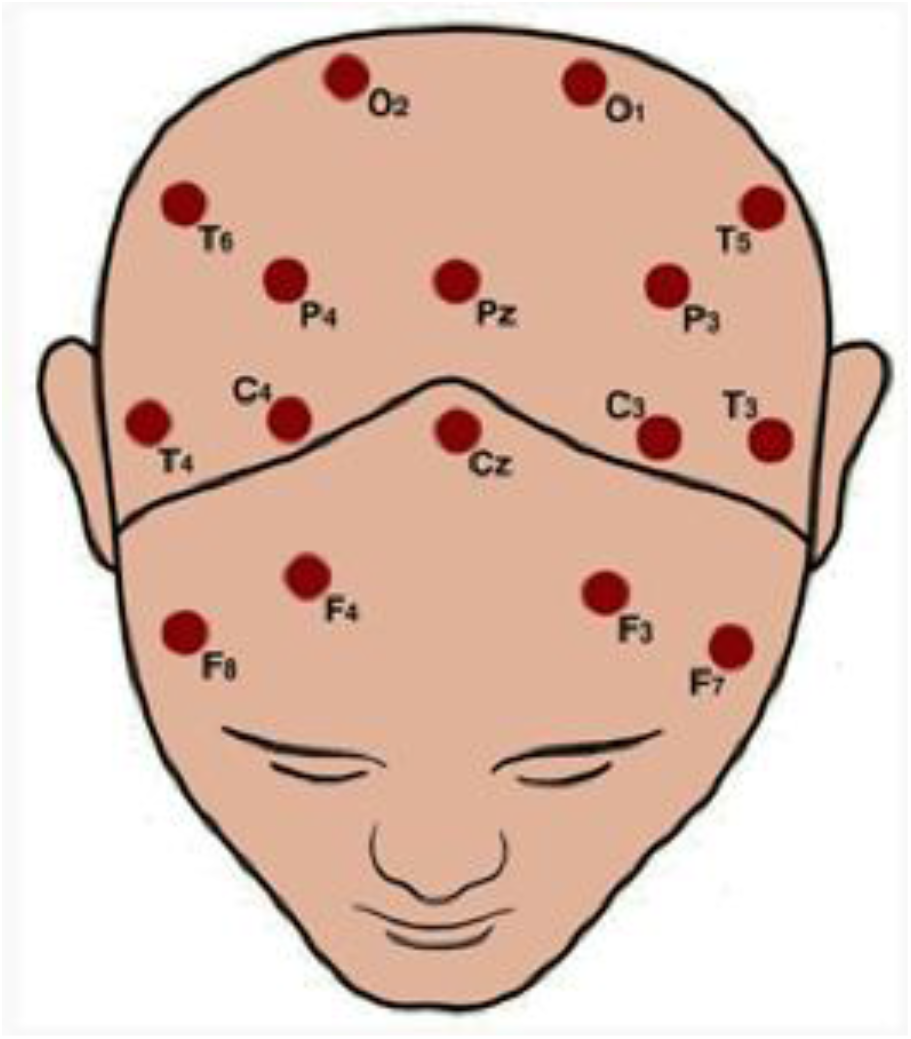
Topology of EEG recording sensors.

The sampling rate is 128 Hz and the recording time was 1 min giving a total of [60 secs × 128] = 7,680 samples. EEG recordings can be downloaded from the website: http://brain.bio.msu.ru/eeg_schizophrenia.htm. EEG time series were re-referenced to the average reference electrode (Nunez and Srinivasan, 2006) before preprocessing steps.

Parents of the participants have given their informed consent for the participation of adolescents to the original study.

Program of experiments has been approved by the local ethical committee of Moscow Mental Health Scientific Center of Russian Academy of medical Sciences.

For further details see the following article (Kulaichev and Gorbachevskaya, 2013).

### 2.1 Artifact Reduction with Independent Component Analysis (ICA) and Wavelets

EEG resting-state activity was corrected for artefacts via a well described procedure. We used the entire 1 min recordings of 7680 time points to the artefact correction pipeline. First, we removed line noise using a notch filter at 50 Hz and (Delorme and Makeig, 2004). We adapted independent component analysis (ICA) and the extended Infomax algorithm implemented in EEGLAB (Delorme and Makeig, 2004). The outcome of this procedure is 16 independent components with a characteristic topography and time course. A given independent component (IC) time course from each IC was further decomposed with discrete wavelet transform and Daubechies wavelet filters. Wavelets decomposed every broadband activity of the IC time series into subcomponents with characteristic carrier brain frequency. Wavelet decomposition of IC time course has been realized in 60 temporal segments of 1 sec each in order to capture in more detail the contamination of an artefact. Artefacts could be detected on specific scalp locations contaminating one frequency or in broaden brain locations and contaminating more than one frequency. Every wavelet subcomponent of an IC is classified as real brain activity or artifactual activity that could be: ocular, muscle or cardiac artefacts.

We estimated kurtosis and skewness values in temporal non overlapping windows of 1 sec for every wavelet subcomponent of an IC. The set of 60 values (60 temporal segments of 1 sec = 128 time points) for both metrics was further z-scored and temporal segments of 1s with zscoring values ±2 were classified as artifactual temporal segments (Dimitriadis et al., 2015). Then, we zeroed those particular temporal segments for specific wavelet subcomponents of an IC. Then, cleaned IC time courses were recomposed from their cleaned wavelet subcomponents. Finally, cleaned wavelet-ICA EEG activity was composed by the cleaned IC time courses.

Apart from this automatic pipeline, we visually inspected IC and wavelet based time-courses and the related scalp topography of every IC to further validate the correction. Power spectrum analysis of every EEG time course before and after the correction revealed more pronounced characteristic peaks. The correlation of the original time course with its corrected wavelet+ICA version in the time domain was above 0.75 on average compared to the ICA only corrected time series which was below 0.3 on average. So, wavelet-ICA maintains better the temporal structure of broadband activity.

We additionally validated the proposed scheme by estimating signal-to-noise ratio (SNR). SNR was estimated by dividing the power spectrum in the frequency domain within every frequency range from the denoised time series with the power spectrum of the original noisy time series. The following equation defines the SNR:

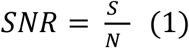

Where S is the power spectrum of the desired cleaned EEG time series and N the power spectrum of the noisy original time series. Before estimating N, we subtracted the power related to the noisy temporal segments detected with our wavelet+ICA method.

We adopted a wavelet-ICA approach for denoising EEG signals with the advantage of not zeroing a whole ICs but only specific temporal segments of 1 sec on wavelet decomposed subcomponents related to specific brain frequencies and extracted via a wavelet transform over each ICs. Figure 1 illustrates the topology of the EEG sensors.

### 2.2 Signal Power Analysis

We estimated the relative power spectrum (RSP) of every EEG sensor and frequency band in both groups. Welch’s algorithm was adopted using MATLAB function pwelch leading to the estimation of power spectrum per frequency band first and then their percentage across total power spectrum leading to RSP. Group statistical analysis across sensors and frequency bands have been performed with Wilcoxon Rank Sum test (p < 0.05, Bonferroni corrected, p’< p/(16*7) where 16 refers to EEG sensors and 7 is the number of the studying frequency bands).

Motivated by a recent study (Howells et al., 2018), we estimated also the ratio of RSP between δ frequency and the two α subbands. Group statistical analysis was performed with Wilcoxon Rank Sum Test (p < 0.05, Bonferroni corrected, p’ < p/(16*2) where 16 refers to EEG sensors and 2 is the number of ratios). Characteristic individual alpha peaks were also estimated from sensor’s spectrum profile of every EEG sensor (Corcoran et al., 2018; see supp.material).

### 2.3 Dynamic imaginary part of phase locking value (iPLV)/ correlation of the envelope (corrEnv) estimates: the Dynamic Integrated Functional Connectivity Graph (DIFCG) based on iPLV and corrEnv (DIFCG^iPLV^/ DIFCG^corrEnv^ graph)

Here, we describe our dominant intrinsic coupling model (DoCM) presented in the majority of functional neuroimaging modalities (Antonakakis et al., 2017a; Dimitriadis, 2017b, 2018c,2018d). The goal of the DoCM model is to extract the dominant coupling mode between every pair of EEG sensors here and across temporal segments. DoCM model defines the dominant coupling mode across intra-frequency phase-to-phase coupling and inter-frequency phase-to-amplitude coupling modes.

We studied dynamic functional connectivity across experimental time and the 16 EEG sensors within and between the seven studying frequency bands {δ, θ, α_1_, α_2_, β_1_, β_2_, γ} defined, respectively, within the ranges {0.5–4 Hz; 4–8 Hz; 8–10 Hz; 10–13 Hz; 13–20 Hz; 20–30 Hz; 30–48 Hz}. EEG recordings were bandpass filtered with a 3rd order zero-phase Butterworth filter using filtfilt.m MATLAB function.

The width of the temporal window was set equal to 250ms (or 32 samples) and moved forward across experimental time every 46 ms (6 samples) which is adequate to capture both fast and slow oscillations (Dimitriadis et al., 2013a,b,2015a,b,2016a,b,c,2017a,b,2018b,c). Within frequency and between-frequency (cross-frequency) interactions between every possible pair of frequencies were estimated for every temporal segment leading to a quasi static Functional Connectivity Graph (FCG). This approach leads to 1275 temporal segments and to a dynamic functional connectivity graph (DFCG).

Here, we adopted two connectivity estimators to quantify within frequency and cross-frequency interactions: the imaginary part of phase locking value (iPLV) and the correlation of the envelope (corrEnv).

To exemplify the notion of cross-frequency coupling estimates to the interested readers that are not familiar, we demonstrated an example between δ and α_1_ frequencies derived from F3 and F4 EEG sensors from the first healthy control subject. Figure 2 demonstrates the algorithmic pipeline for estimating phase-to-amplitude coupling (PAC) with iPLV estimator. Figure 2A,B illustrate the time course of the first 5 secs from the F3 and F4 EEG sensor activity, correspondingly. Figure 2C demonstrates the time course of δ activity from the F3 sensor while Figure 2D shows the time source of α_1_ activity from the F4 sensor and the δ activity extracted from α_1_ activity from the same sensor. δ activity was extracted from α_1_ activity using the bandpass filters transfer function coefficients used to get δ activity from the F3 sensor. Figure 2E shows the δ activity within α_1_ activity from the F4 sensor while Figure 2F shows its phase temporal course extracted via Hilbert transform. Phase time course of δ activity from F3 sensor is illustrated in Figure 2G while both targeted phase time series are shown in Figure 2G. Last sub-figure 2I presents the phase difference of the two targeted phase time series from which iPLV will quantify PAC strength. Intra-frequency coupling with iPLV is estimated using the phase difference time series as shown in Figure 2I derived from two time series with the same frequency content. The whole procedure is repeated for 21 cross-frequency coupling estimates for both CFC estimators and in a dynamic fashion. The 7 within frequency couplings have been estimated between the phase temporal course from two times series with the same frequency content using iPLV.

**Figure 2.**
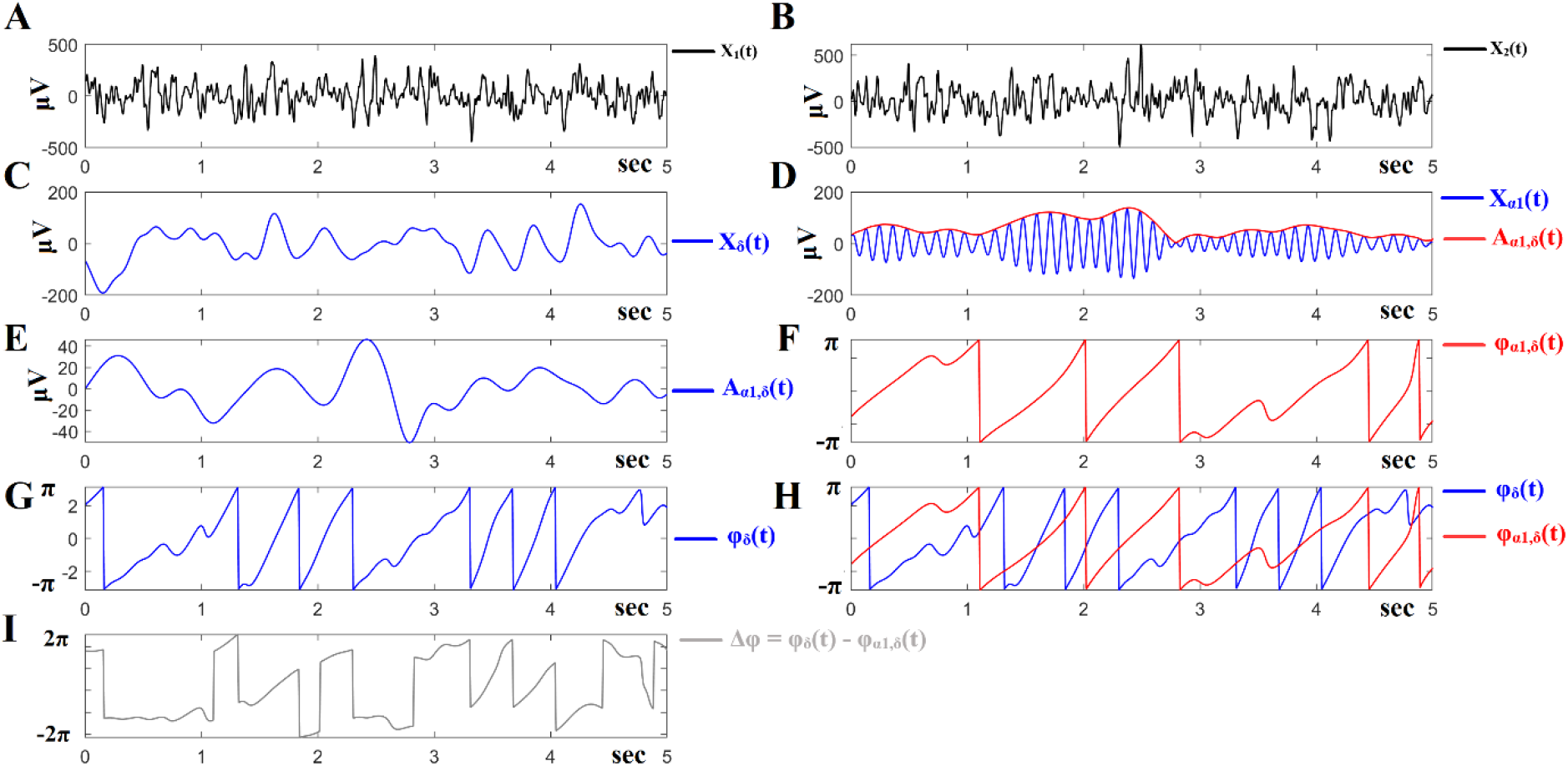
Outline of the PAC algorithm. A,B illustrate the time course of the first 5 secs from the F3 and F4 EEG sensor activity. C demonstrates the time course of δ activity from the F3 sensor while D shows the time source of α_1_ activity from the F4 sensor and the δ activity extracted from α_1_ activity from the same sensor. δ activity was extracted from α_1_ activity using the bandpass filtering transfer functions employed to extract δ activity from the F3 sensor. E shows the δ activity within α_1_ activity from the F4 sensor while Figure F shows its phase temporal course extracted via Hilbert transform. Phase time course of δ activity from F3 sensor is illustrated in G. Both final targeted phase time series are shown in G. I presents the phase difference of the two targeted phase time series from PAC strength will be quantified via iPLV estimator.

Figure 3 is devoted to describe the algorithmic steps of amplitude-to-amplitude coupling (AAC) between pairs of frequencies as a second complementary cross-frequency coupling estimator. Figure 3A,B refer to the same time courses as in Figure2A,B from the first 5 secs from F3 and F4 EEG sensors. Figure 3C illustrates the δ activity from the F3 sensor and its related envelope extracted via Hilbert transform. Figure 3D illustrates the α_1_ activity from the F4 sensor and its related envelope extracted via Hilbert transform. Figure 3E and F show the envelopes from F3 and F4 sensors, correspondingly while Figure 3G demonstrates both envelopes in a common plot. AAC is estimated between those time courses via a correlation envelope analysis (corrEnv). Intra-frequency coupling with corrEnv is estimated between pairs of envelopes extracted from time series with the same frequency content. The whole procedure is repeated for 21 cross-frequency coupling estimates for both CFC estimators and in a dynamic fashion. The 7 within frequency couplings have been estimated between the envelope temporal course from two time series with the same frequency content using corrEnv.

**Figure 3.**
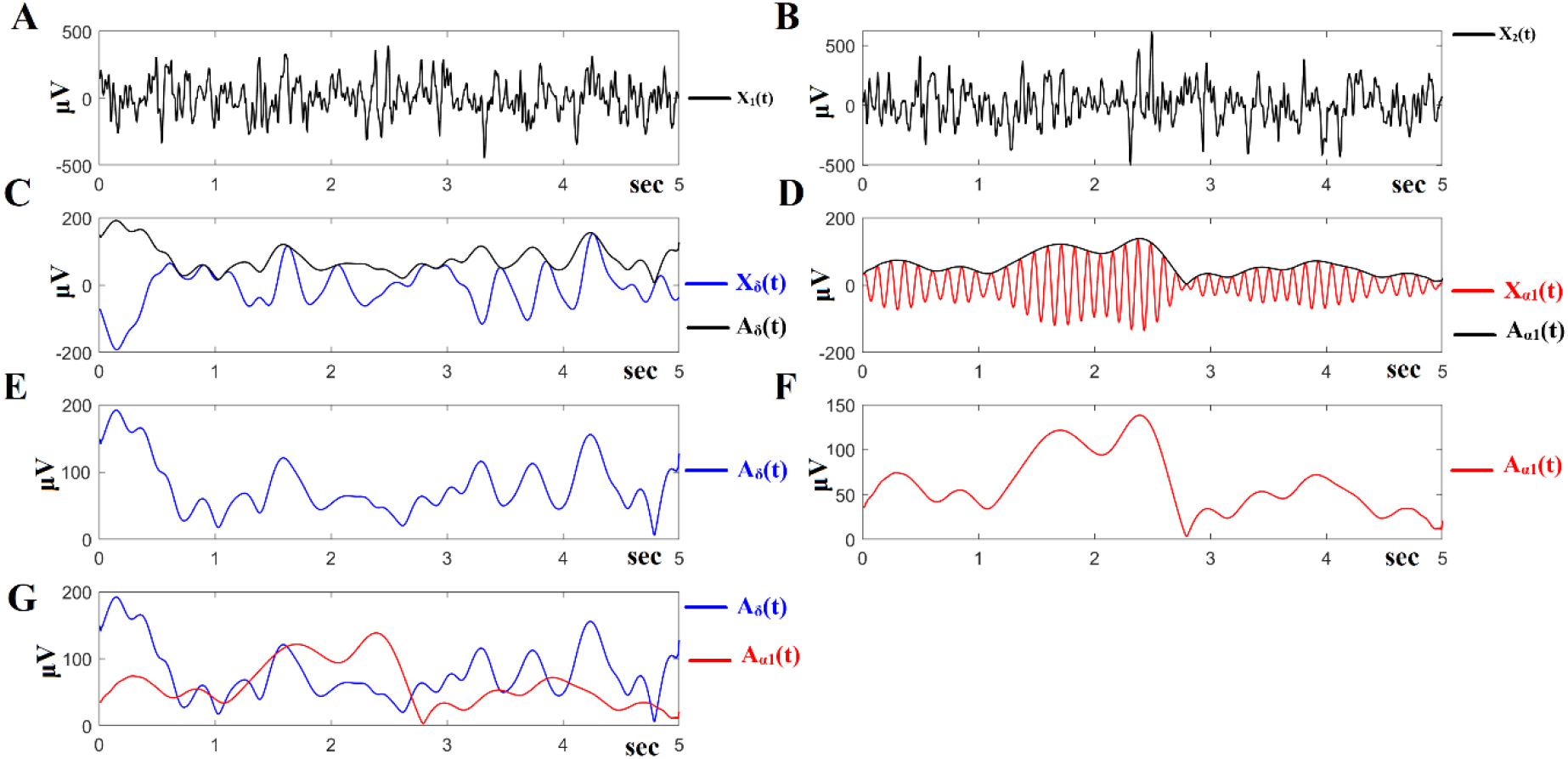
Outline of AAC algorithm. A,B show the same time courses as in Figure2A,B from the first 5 secs from F3 and F4 EEG sensors. C illustrates the δ activity from the F3 sensor and its related envelope extracted via Hilbert transform. D demonstrates the α_1_ activity from the F4 sensor and its related envelope extracted via Hilbert transform. E and F show the envelopes related to δ activity of F3 and α_1_ activity of F4 sensor. G demonstrates both envelopes in a common plot. AAC is estimated between those time courses via a correlation envelope analysis (corrEnv).

This procedure which is described in detail in our previous papers and also aforementioned (Dimitriadis and Salis, 2017b; Dimitriadis et al., 2018c), resulted in 7 DFCG^iPLV^/ 7 DFCG^corrEnv^ per participant for within frequency bands and 21 DFCG^iPLV^/21 DFCG^corrEnv^ per participant for each possible cross-frequency pair. 7 DFCG^iPLV^/ 7 DIFCG^corrEnv^ tabulate iPLV/corrEnv estimates between every possible pair of sensors. For each subject, two 4D tensors [frequencies bands (28 × temporal segments × sensors × sources] were constructed for each subject integrating spatiotemporal phase and amplitude based interactions. The final outcome of this analysis is two 4D fully-weighted tensors. However, not all the connections exist. For that reason, we adapted a proper surrogate analysis.

The adopted surrogate analysis focuses on detect ‘true’ brain interactions between every pair of sensors and at every temporal segment (snapshot of DFCG) and finally to detect the dominant type of interaction. For this purpose, we constructed 1,000 surrogate time-series by cutting first at a single point at a random location around the middle of the original time series (between 25 s and 35 s), creating two temporal segments and then exchanging the two resulting temporal segments (Dimitriadis and Salis, 2017b; Dimitriadis et al., 2018c). Surrogate time series were created on the 7 bandpass filtering time series in the amplitude domain and were employed for both connectivity estimators per subject.

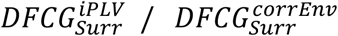 were estimated over the 1,000 surrogate time series for both within frequencies and between frequencies interactions, for each pair of sensors and for each temporal segment. 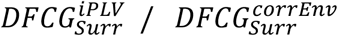 are 5D tensors where the 1^st^ dimension refers to 1,000 surrogate values while the rest 2^nd^ −5^th^ dimensions are similar to the 4D tensors of the original DFCG^iPLV^/ DFCG^corrEnv^. Comparing the original value for every EEG sensor pair, for every temporal segment and every coupling mode (28 in total) from the original DFCG^iPLV^/ DFCG^corrEnv^ with the 1,000 surrogates (1^st^ dimension of 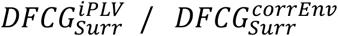), we assigned a p-value to every potential intrinsic coupling mode. We then correct for multiple comparisons the p-values related to the 28 (21 + 7) possible DoCM in order to reveal a DoCM per pair of EEG sensors and for each temporal segment. There are three scenarios:

1. a no *p*-value survived the multiple correction (*pʹ* < p/28) where *p* = 0.05)
2. we selected the DoCM with the maximum iPLV/corrEnv value if more than one survived,
3. only one coupling mode survived the multiple correction

Figure 4A demonstrates how DoCM are defined employing the first two temporal segments from a healthy control subject between F7 and F3 EEG sensors. The surrogate-based analysis revealed α_2_-γ phase-to-amplitude coupling (PAC) as the dominant coupling mode (DoCM) among the 28 potential coupling modes for the two very first temporal segments between F3-F7 sensors. Matrices called comodulograms tabulate the strength measured with iPLV for within-frequency couplings (main diagonal) and between-frequencies (off diagonal). Figure 4B illustrates the fluctuation of DoCM across experimental time for F3-F7 sensor pair. Y-axis refers to one of the 28 potential coupling modes while the colour refers to the iPLV related strength. Coloured matrix on the right tabulated the probability distributions (PD) of every potential coupling modes across experimental time. α1-α_2_ is the CFC with the highest representation across experimental time for this EEG pair of sensors. Flexibility Index (FI) and PD are estimated from semantic time series as the one presented in Figure 4B for every pair of EEG sensors (see next section).

**Figure 4.**
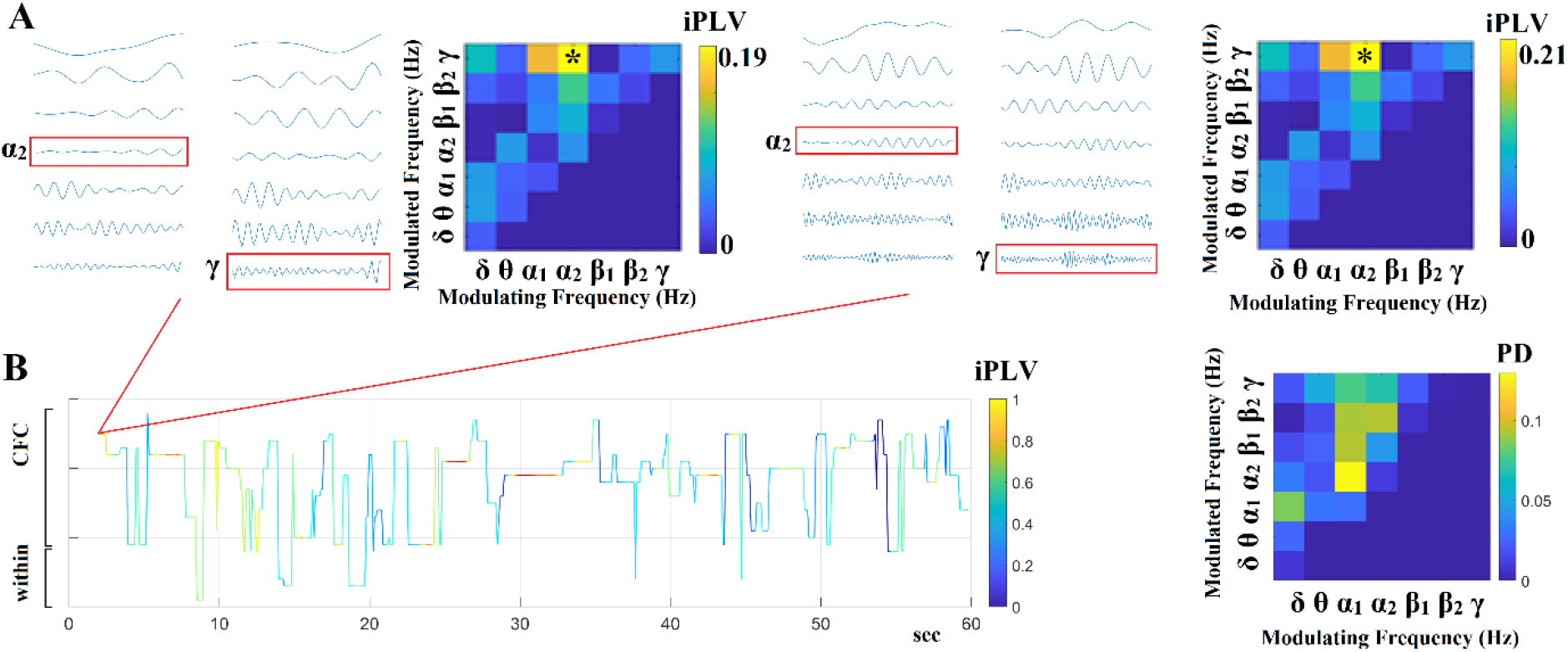
Determining Dominant Intrinsic Coupling Modes (DoCM) based on iPLV. (A) A detailed schematic illustration of the adapted DoCM model showing the detection of dominant coupling mode between two EEG sensors (F3 and F7) for the first two consecutive temporal segment (ts_1_, ts_2_). For demonstration purpose, we adapted imaginary Phase Locking (iPLV) which was employed for the estimation of both within frequencies (e.g., δ to δ) and between frequencies (cross-frequency) interactions (e.g., δ to θ). Surrogate analysis will reveal the DoCM for both temporal segments. During ts_1_ the DoCM reflected significant phase locking between α_2_ and γ oscillations (indicated by red rectangles) while during in ts_2_ the dominant interaction was remain stable. (B) Burst of DoCM between the F3 and F7 sensors. This streaming of the basic elements of neural communication is encapsulated in the DoCM time series to form a neural “word.”, which can be interpreted as a spatio-temporal message in the macroscale level (Buzsaki & Watson, 2012). The plot illustrates the fluctuation of DoCM across experimental time for F3-F7 sensor pair. Y-axis refers to one of the 28 potential coupling modes while the colour refers to the iPLV related strength. The first two temporal segment showed in A. revealed α_2_ – γ as DoCM. This observation is shown in the first samples of the time-source showed in B. FI is estimated based on such a semantic time series by counting how many times the DoCM changed between consecutive temporal segments divided by the total number of temporal segments −1. FI ranged within [0,1] where higher values are interpreted as higher flexibility. The comodulogram on the right demonstrates the probability distribution (PD) of the DoCM across temporal segments for the EEG sensor pair F7-F3. α_1_-α_2_ is the CFC with the highest representation across experimental time for this EEG pair of sensors The outcome of this approach is the construction of 2 DFCG represented as 3D tensors of size [1275×16×16].

After applying a multiple comparison correction, the strength and also the type of dominant coupling mode are tabulated in 2 3D tensors of size [1275 × 16 × 16] called Dynamic Integrated Functional Connectivity Graph (DIFCG). We used the term integrated to underline the information of DoCM tabulated within these DIFCGs. One keeps the strength of the coupling based on iPLV/corrEnv and the second tabulated the dominant coupling mode of interaction using integers from 1 up to 28:1 for δ, 2 for θ, …, 7 for γ, 8 for δ − θ, …, 28 for β_2_ − γ} (Figure 4B). We followed the same procedure in the previous studies from our group (Dimitriadis and Salis, 2017b, Dimitriadis et al., 2017c,2018a,b,c,d).

Figure 4B demonstrates the temporal evolution of DoCM (1^st^ 3D tensor) and the related strength (2^nd^ 3D tensor) across experimental time for F7-F3 EEG pair while the comodulogram on the right side tabulates the PD of the DoCM for this semantic time series. FI and PD will be estimated over the 2^nd^ semantic 3D tensor.

In the present study, we adapted the debiased version of PAC as presented in van Driel et al., (2015).

### 2.4 Semantic Features Derived from the evolution of DoCM

This section describes the semantic features that can be extracted by analysing the 2^nd^ 3D tensor that preserves the DoCM across spatio-temporal dimensions.

#### 2.4.1 Flexibility Index (FI)

We adopted a previously defined estimator called Flexibility index (FI) which quantifies the transition rate of DoCM between every pair of sensors across experimental time (Dimitriadis and Salis,2017b; Dimitriadis,2018c). FI is estimated based on the 2^nd^ 3D tensor of the DIFCG that tabulates the semantic information of DoCM across the brain and experimental time. This metric will called hereafter FI^DoCM^ which is defined as:

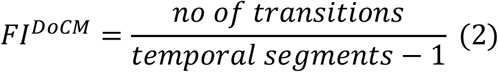

FI^DoCM^ gets higher values for higher “jumps” of DoCM between a pair of EEG sensors across experimental time. Figure 4B illustrates how FI^DoCM^ is estimated for F3-F7 EEG pair. This approach leads to a 16 × 16 matrix per subject or 16^2^ =140 FI features. We estimated also the global FI by averaging the 16 nodal FI. FI is estimated from a semantic time series presented in Figure 4B.

#### 2.4.2 Spatiotemporal distribution of DoCM— Comodulograms

Based on the 2^nd^ 3D DIFCG that keeps the semantic information of the preferred dominant coupling mode, we can tabulate in a frequencies × frequencies matrix the probability distribution (PD) of observing each of the DoCM frequencies across 7 (intra-frequency) + 21 (cross-frequency coupling) = 28 possible coupling modes and exploring their spatio-temporal distribution.

The spatiotemporal PD tabulated in a matrix is called hereafter comodulogram and an example is demonstrated in Figure 4B (Antonakakis et al., 2016,2017a,b; Dimitriadis and Salis,2017b,c; Dimitriadis,2018a,c,d). PD is a new vector of features with dimension 1 × 28 possible DoCM. Figure 4B visualizes a PD matrix and demonstrates how it is computed.

#### 2.4.3 Nonlinearity Index (NI) based on DoCM

When two brain areas communicate via within the same frequency (intra) then this interaction is linear. However, cross-frequency interactions are nonlinear pathways of information exchange via brain areas. We first estimated a global PD of dominant coupling modes across space (EEG pairs) and time (temporal segments). This calculation yields a matrix of size 28 (potential coupling modes) × 1275 (temporal segments) per subject. From this global PD matrix, we estimated the sum of PD related to cross-frequency coupling (21 values) versus the sum of PD related to within frequency coupling (7 values). This ratio can serve as a nonlinearity index (NI) of the information flow between brain areas (equation 3). To untangle the role of α modulated frequency as a key driver of any significant group differences, we also estimated the ratio of the sum of PD of cross-frequency couplings between a modulated frequency and the rest of frequencies versus the PD of the within-frequency interactions on the modulated frequency was estimated. Particularly, we estimated the ratio of the sum of PDs between α_1_ and {α_2_,β_1_,β_2_,γ} and between α_2_ and {β_1_,β_2_,γ} with the sum of PDs related to α_1_ and α_2_ within frequencies interactions (equation 4). The NI gets a positive value where the higher it gets the higher is the contribution of CFC to the dominant coupling modes and so the higher the nonlinearity of the communication within the complex system. For group comparisons, we adopted Wilcoxon Rank-Sum test as a statistical test to compare the distributions of temporal NI.

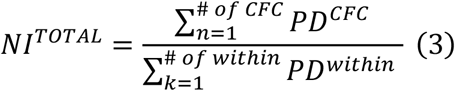

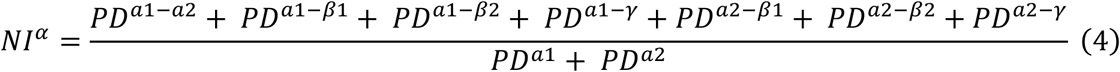

### 2.5 Inter-Hemispheric Group Differences of Dynamic Functional Connectivity Strength

We explored how inter-hemispheric dynamic functional strength differed between the two groups. Temporal functional connectivity strength is a time series of size 1275 that informs us how functional connectivity strength changes over experimental time. Figure 5A illustrates an example of such a times series for iPLV estimator between F7-F8 EEG sensors from the first healthy control subject. Our analysis focused on functional connectivity strength between the following two sets of EEG sensors, one per hemisphere: Left hemisphere: {O1 P3 T5 C3 T3 F3 F7}, right hemisphere: {O2 P4 T6 C4 T4 F4 F8}. The combination of both sets forms a set of 49 inter-hemispheric links. From every functional connectivity strength time series, we estimated the mean value and also its spectrum using Welch’s approach and the related function in MATLAB. For the spectrogram, we extracted the dominant frequency that is a descriptive statistic of the fluctuation of connectivity across experimental time (Figure 5B).

**Figure 5.**
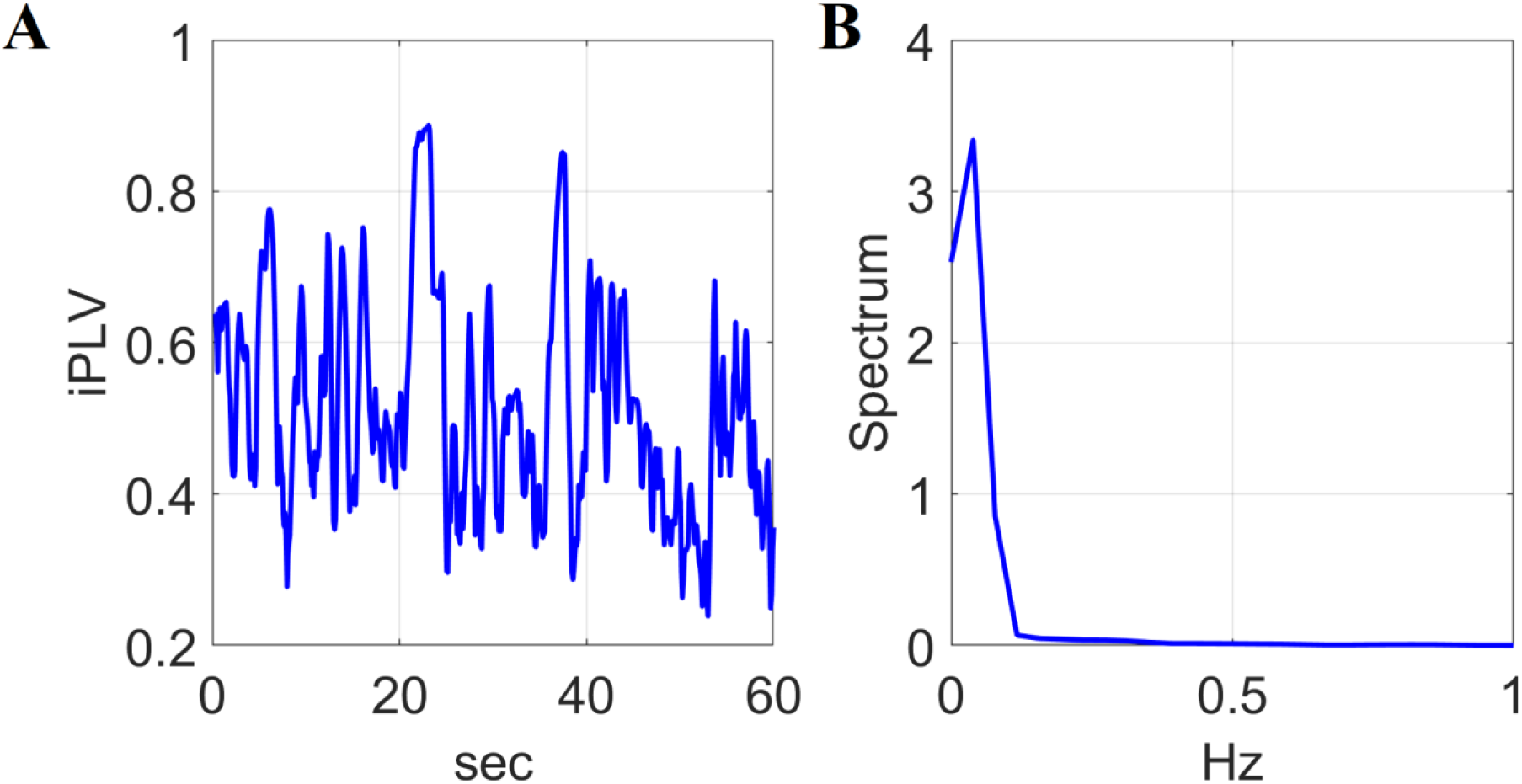
Inter-hemispheric fluctuations and the related dominant frequency A illustrates an example of a time series presenting the temporal functional strength between F7-F8 EEG sensors from the first healthy control subject. The spectrogram of this dynamic functional strength was extracted and the related dominant frequency is assigned to this inter-hemispheric pair as descriptive statistic of the fluctuation of connectivity across experimental time (B).

Group statistical analysis was performed with Wilcoxon Rank Sum Test (p < 0.05, Bonferroni corrected, p’ < p/49 where 49 refers to the total number of inter-hemispheric links).

### 2.6 Group Discrimination via a Machine Learning Approach

We adopted the simplest k-NN classifier in order to enhance the quality of the features extracted with our DoCM model. PD and FI were integrated into a single feature vector of 284 features (28: PD + 256 (16×16): FI).

We classified every subject with k-NN classifier (Knn = 10) following a 5-fold cross-validation (CV) scheme running 100 times. As a feature selection algorithm, the infinite feature selection algorithm was selected (Roffo et al., 2015). Feature selection has been applied within every fold of the 5 CV and across runs. From the ranking of 284 features within every fold, we kept the first 10 ranked features. We scored the features that were selected in the first 10^th^ across the 100 runs and 5 folds.

Our analysis was followed independently for iPLV/corrEnv derived PD and FI.

### 2.7 Software

The analysis has been realized in MATLAB environment (v2019b) using the signal processing toolbox. Fast ICA has been adopted from the fieldtrip toolbox. Dynamic functional connectivity has been based on in-house software provided in our github website: https://github.com/stdimitr/time_varying_PAC.

## 3. Results

Our results reported on cleaned EEG brain activity with a combination of ICA and wavelets. SNR values were higher than 6 while no group differences have been estimated at single-channel comparisons (SFigure 1). The high SNR values further support current findings under the framework of connectivity analysis which is affected by SNR.

### 3.1 Relative Signal Power in HC and SSDs subjects

Statistical comparison of RSP between the two groups across EEG sensor space and frequency bands revealed significantly higher signal power in δ (increased synchronization) and also in θ frequency bands for SSDs compared to HC evaluating our first hypothesis. We found also significant lower RSP values for the SSD group compared to HC in the α_2_-frequency band supporting our second hypothesis of reduced alpha desynchronization in SSDs subjects.

Figure 6 illustrates group-averaged RSP in SSDs and HC group. Comparison of group-averaged Relative Signal Power (RSP) of δ versus RSP of α_1_ and α_2_ across EEG sensor space in HC and SSDS patients revealed interesting trends. Our analysis revealed significantly higher ratio RSP of δ / α_1_ for SSDS compared to HC in F7 and Cz EEG sensors. Significantly higher ratio RSP of δ / α_2_ for SSDS compared to HC was revealed in all the EEG sensors with the exception of channel T6 (Figure 7) (* Wilcoxon Rank Sum test (p < 0.01, Bonferroni corrected, p’< p/(16*2) where 16 refers to EEG sensors and 2 to the number of the ratios). These results supported a distinct role of alpha subbands.

**Figure 6.**
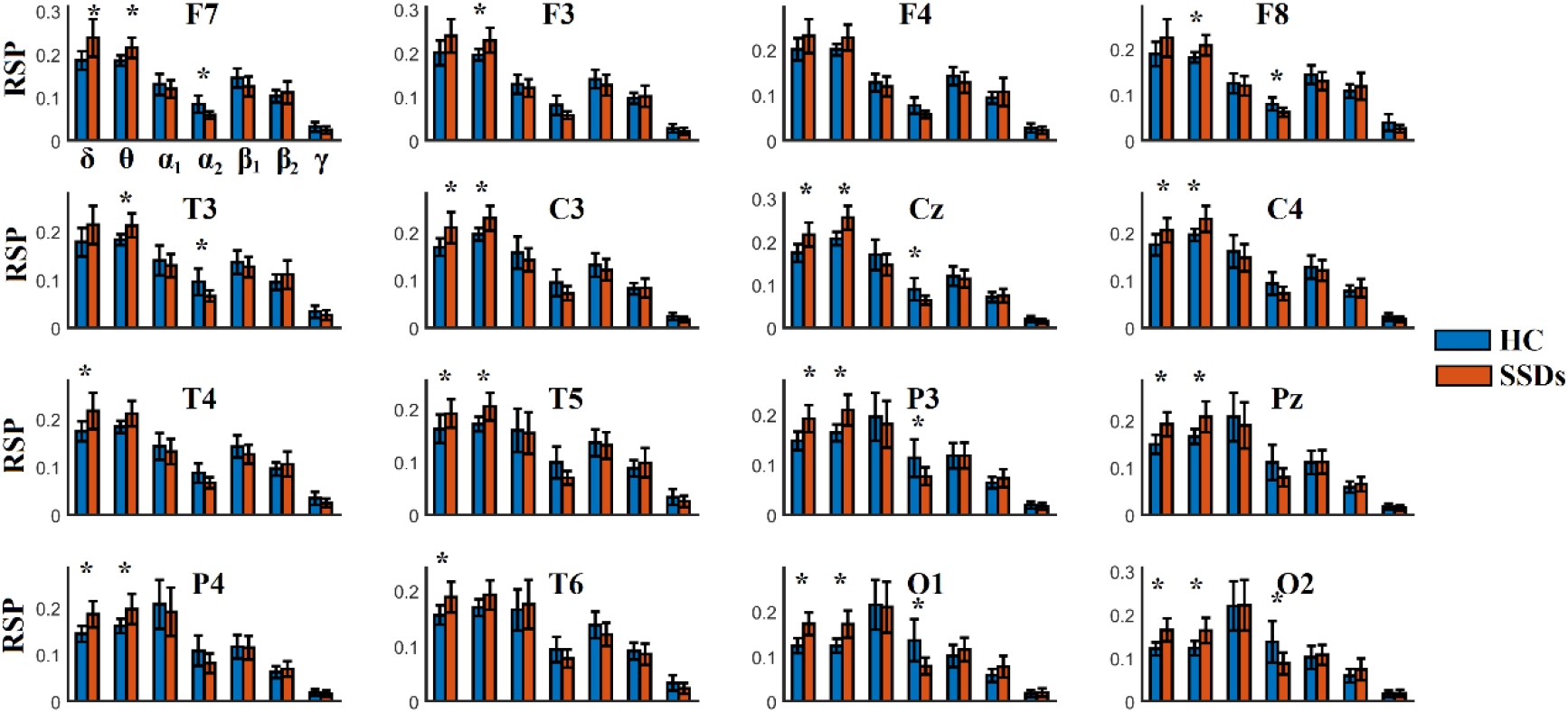
Group-Averaged Relative Signal Power (RSP) across EEG sensor space and frequency bands in HC and SSDS patients. Our analysis revealed significant higher δ RSP for SSDs compared to HC in O1 and O2 EEG sensors. (* Wilcoxon Rank Sum test (p < 0.01, Bonferroni corrected, p’< p/(16*7) where 16 refers to EEG sensors and 7 to the number of the studying frequency bands).

**Figure 7.**
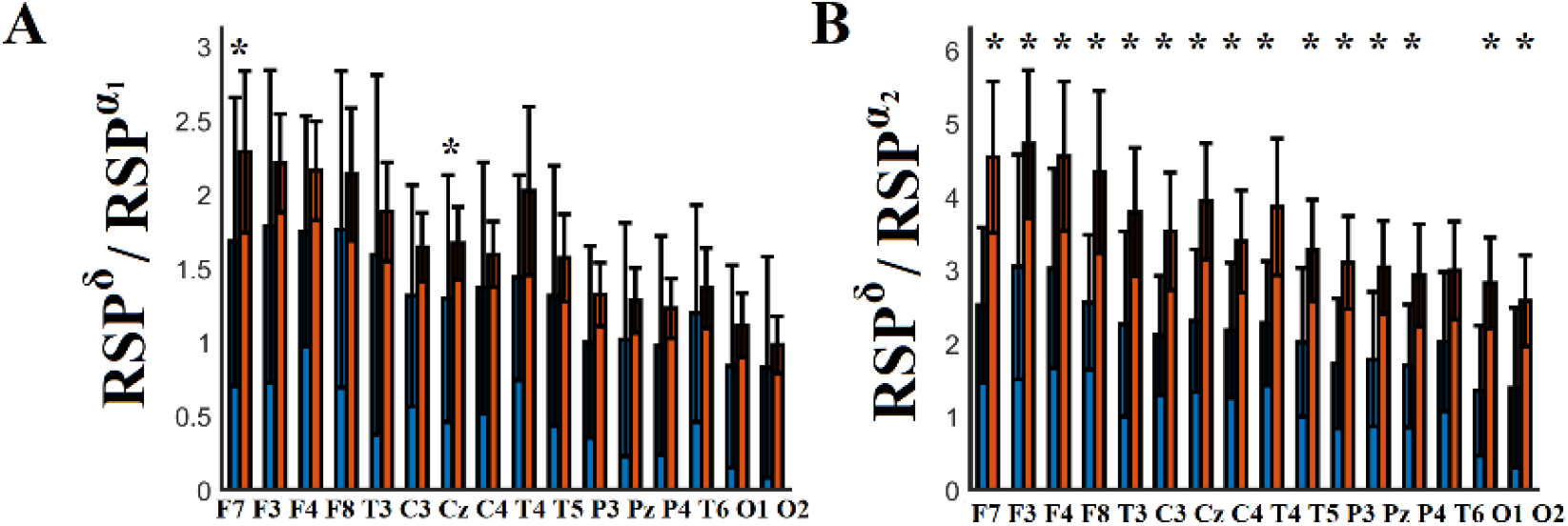
Group-averaged Ratio of Relative Signal Power (RSP) of δ versus RSP of α_1_ and α_2_ across EEG sensor space in HC and SSDs patients. Our analysis revealed significant higher ratio of RSP of **δ with α_1_** for SSDS compared to HC in F7 and Cz EEG sensors. Significant higher ratio RSP of **δ with α_2_** for SSDS compared to HC were revealed in all the EEG sensors with the exception of T6 EEG sensors. (* Wilcoxon Rank Sum test (p < 0.01, Bonferroni corrected, p’< p/(16*2) where 16 refers to EEG sensors and 2 to the number of the ratios).

Group-averaged alpha peaks was around 9.03 Hz across EEG sensors. Alpha peaks didn’t reveal any group differences trends (SFigure 2).

### 3.2 Contribution of Semantic Features based on DoCM model for High Classification Performance of SSDs Subjects

Our machine learning strategy revealed 10 consistent features among FI and PD of DoCM in both estimators. Consistent features refer to those features where their Score across 100 runs × 5 – fold CV equal to 500. Figure 8 (a-b) illustrates the spatial distribution of the FI across every possible pair of EEG sensors for HC and SSDS group while Figure 8 (c-d) illustrates the comodulograms of the PD for HC and SSDs group tailored to iPLV. Similarly, Figure 9 (a-b) illustrates the spatial distribution of the FI across every possible pair of EEG sensors for HC and SSDs group while Figure 9 (c-d) illustrates the comodulograms of the PD for HC and SSDs group tailored to corrEnv. Interestingly, the most discriminative features were the ones related to PD compared to FI.

**Figure 8.**
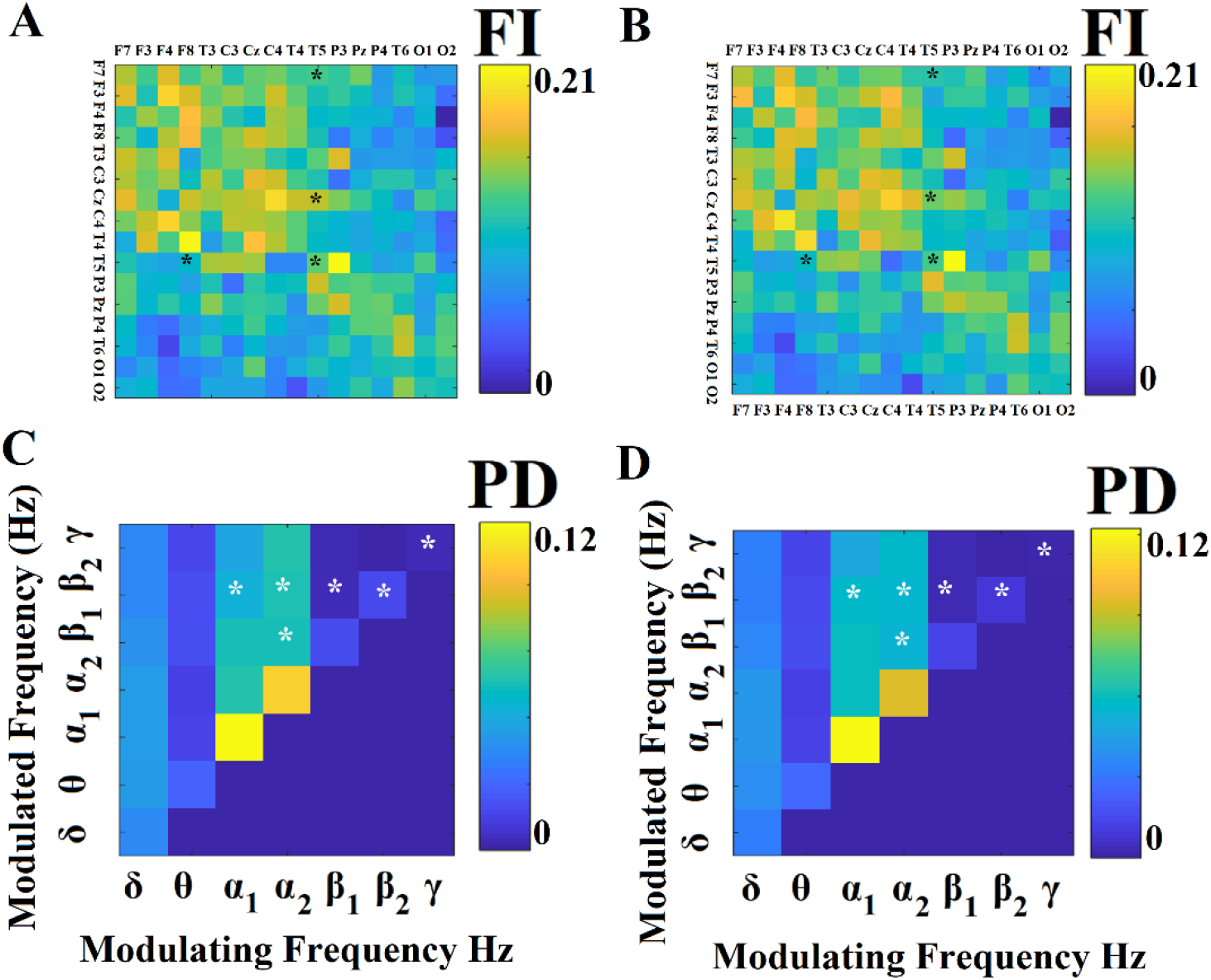
Group-Averaged Flexibility Index (FI) and Comodulograms that tabulate the PD of spatio-temporally DoCM based on iPLV estimator. (A-B) Group-averaged FI for healthy control (HC) group (A) and group with schizophrenia-spectrum disorders (SSDs) (B). (C-D) Group-averaged comodulograms for HC and SSDs group, correspondingly Every FI and PD has been selected via the feature selection algorithm is denoted with ‘*’.

**Figure 9.**
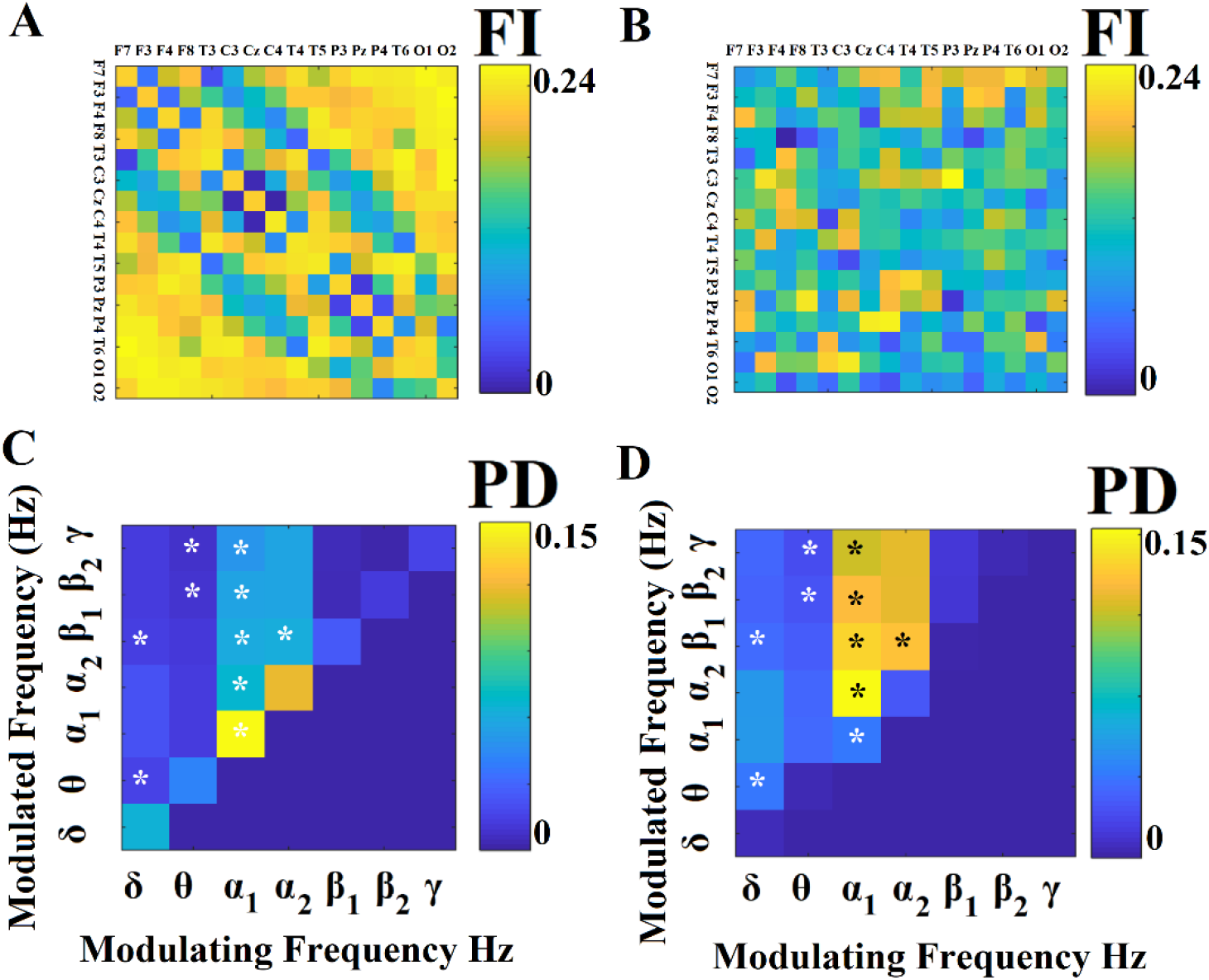
Group-Averaged Flexibility Index (FI) and Comodulograms that tabulate the PD of spatio-temporally DoCM based on corrEnv estimator. (A-B) Group-averaged FI for healthy control (HC) group (A) and group with schizophrenia-spectrum disorders (SSDs) (B). (C-D) Group-averaged comodulograms for HC and SSDs group, correspondingly Every FI and PD has been selected via the feature selection algorithm is denoted with ‘*’.

Figure 10 illustrates the most discriminative features which are PD for θ-γ, α_1_-α_2_ θ-β_2_ crossfrequency pairs. Every dot refers to a subject presented in a 3D feature space. Based on our findings, we evaluated hypothesis III.

The classification performance based on selected FI+PD features was 74% for iPLV and 100% for corrEnv. The weighed discriminative score of features related to corrEnv was 10 times higher than the features from iPLV which was reflected to the absolute accuracy. Moreover, Figure 10 illustrates and evaluates the high classification accuracy between the two groups.

**Figure 10.**
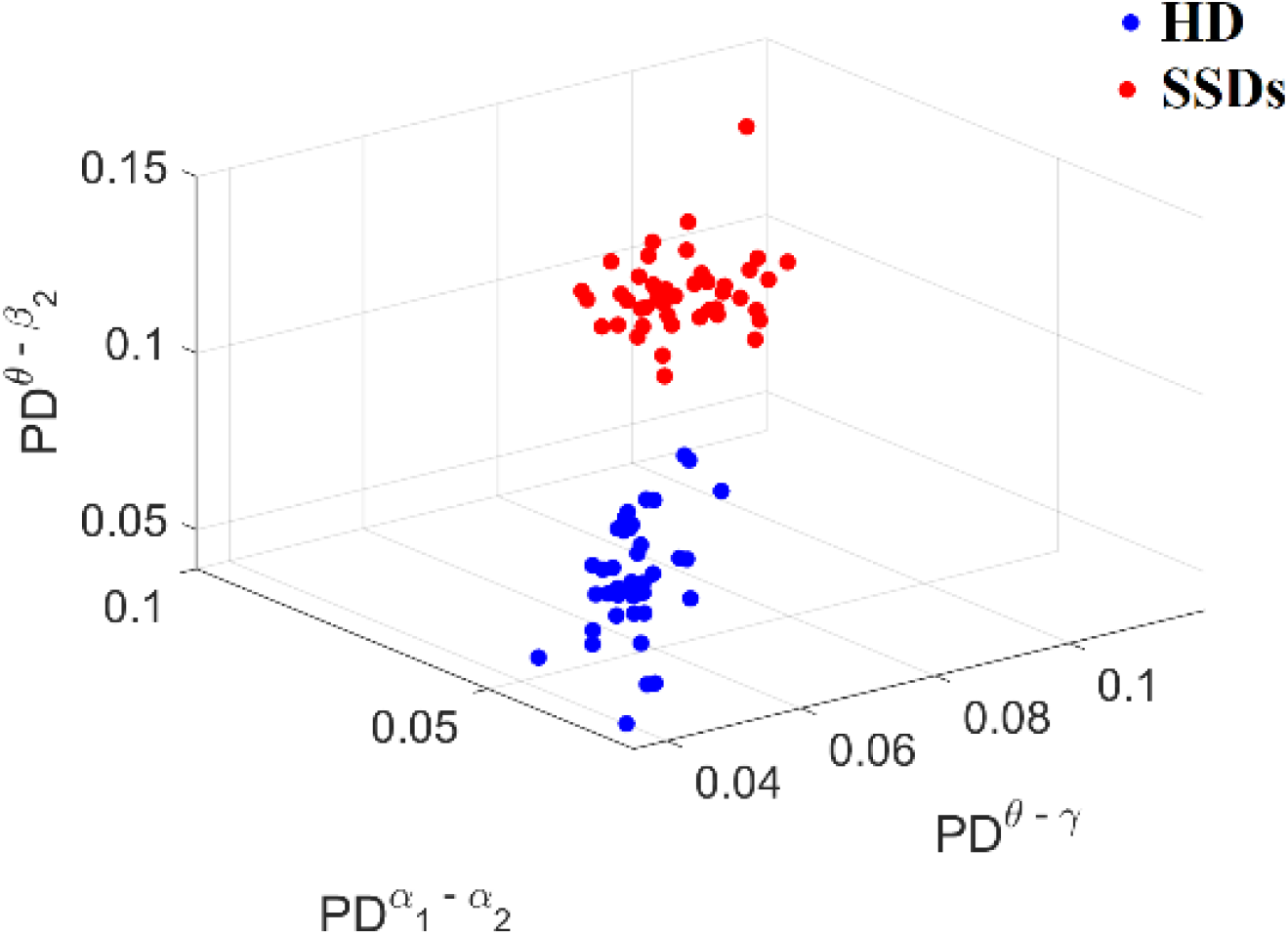
Discriminative power of PD for θ-γ, α_1_-α_2_ θ-β_2_ cross-frequency pairs. Every dot corresponds to a single subject.

Table 1 tabulates the performance for each set of features.

**Table 1.** Evaluation of Classification performance based the semantic features pool of PD and FI and on a multi - kernel SVM approach.

|  | **Accuracy** | **Sensitivity** | **Specificity** |
| --- | --- | --- | --- |
| **PD+FI for iPLV** | 74.53 ± 2.11 | 72.21 ± 2.31 | 71.85 ± 1.89 |
| **PD+FI for corrEnv** | 100.00 ± 0.00 | 100.00 ± 0.00 | 100.00 ± 0.00 |

### 3.3 Global Network Flexibility Index (FI)

By averaging the FI values across both dimensions of the matrix [16 × 16] per subject, we estimated the global network FI as a unique characterization of network flexibility. Group-averaged FI did not reveal significant differences for the iPLV estimator (FI^HC^ = 0.2104, FI^SSDs^ = 0.2085,pval = 0.0673). However, group-averaged FI revealed significant differences for corrEnv estimator (FI^HC^ = 0.2466, FI^SSDs^ = 0.2123,pval = 0.00345 × 10^−5^). Our results supported our hypothesis IV.

### 3.4 Nonlinearity Index (NI) of Information Flow based on DoCM

Figure 11 demonstrates the group-averaged Nonlinearity Index for iPLV (Figure 11.A) and corrEnv estimator (Figure 11.B). It is clear that NI for corrEnv estimator was significant higher for SSDs compared to HC following a Wilcoxon Rank Sum Test across experimental time (p = 0.0001274 × 10^−12^). In contrast, the Nonlinearity Index did not differ between the two groups for iPLV (p = 0.0669). Statistical analysis has been applied over temporal mean of NI time series.

**Figure 11.**
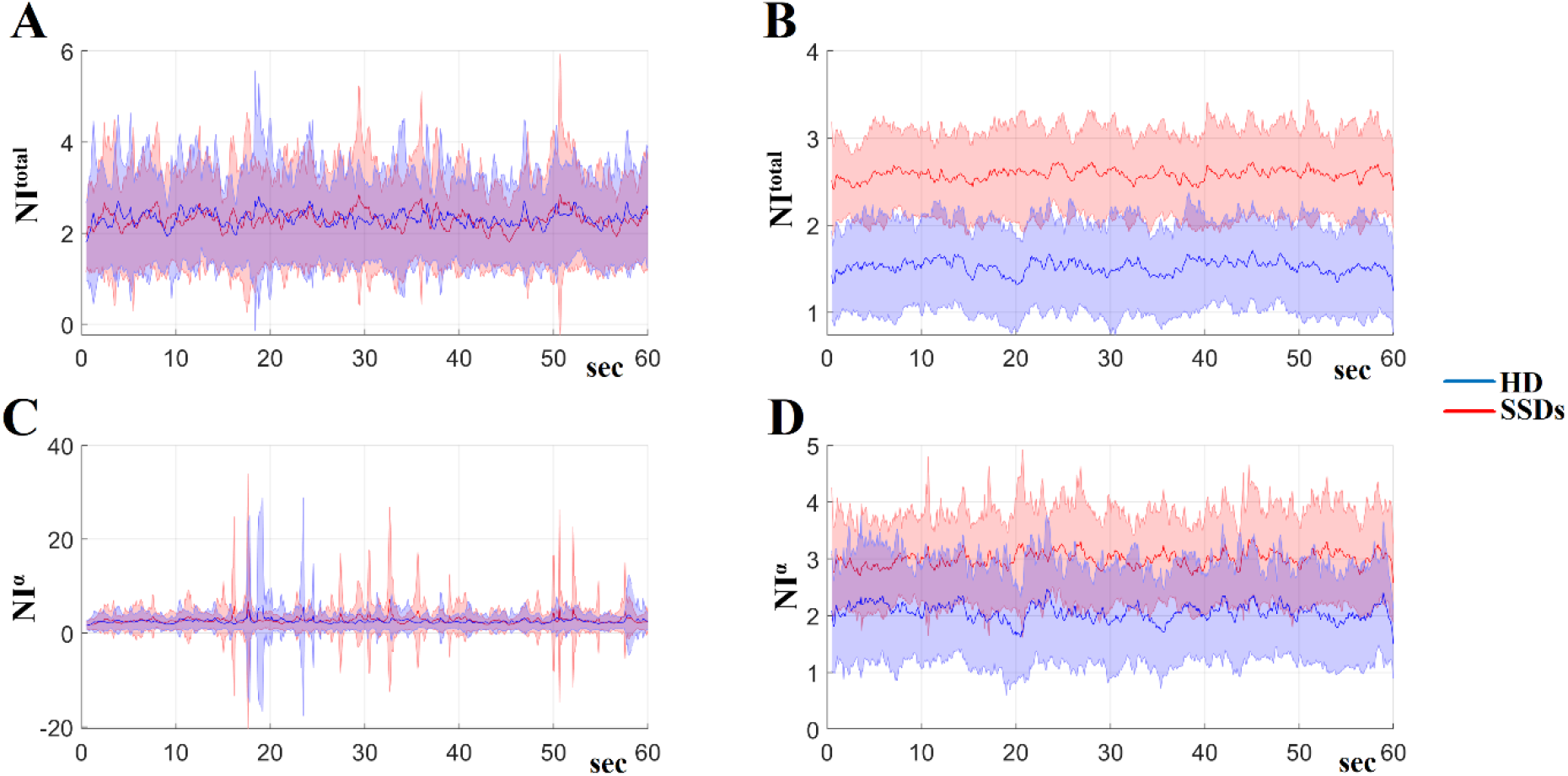
Group –averaged Nonlinearity Index across experimental time. A) NI^Total^ for iPLV, B) NI^Total^ for corrEnv, C) NI^α^ for iPLV D) NI^α^ for corrEnv

### 3.5 Distinct Role of Amplitude and Phase DoCM

Amplitude and phase-based DoCM showed a distinct role in both groups. Both PD of DoCM (Figure 8 and 9) and Nonlinearity Index (Figure 11) supported our hypothesis V. Phase-based intrinsic coupling modes are more sensitive in disorders with functional alterations while amplitude-based intrinsic coupling modes are more sensitive in the presence of a predominant structural alteration (Engel et al., 2013). In a recent study, they highlighted that cortical phase and amplitude coupling patterns shared complementary information revealing the importance of amplitude coupling measures (Siems and Siegel, 2020).

### 3.6 Inter-Hemispheric Group Differences of Temporal Functional Connectivity Strength

No group differences have been identified for the dominant frequency of dynamic functional connectivity strength of inter-hemispheric links with both connectivity estimators. However, we found group-averaged differences between the mean values of the following inter-hemispheric links:O1-P4 P3-O2 P3-T6 C3-T6 F3-O2 F3-C4 (Figure 12.A), for iPLV and C3-T6 C3-T4 for corrEnv (Figure 12.B). Group-averaged iPLV values were higher for SSDs while group-averaged corrEnv were higher for healthy controls.

**Figure 12.**
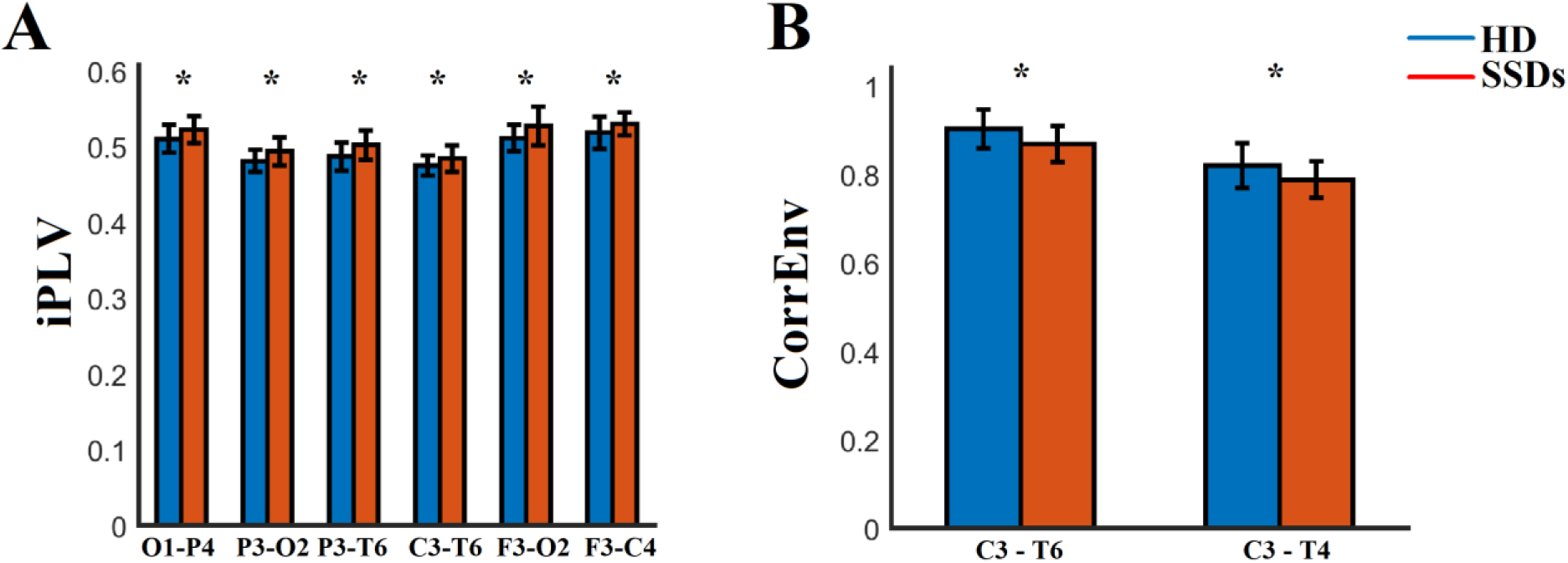
We found group-averaged differences between the mean values of the following inter-hemispheric links:O1-P4 P3-O2 P3-T6 C3-T6 F3-O2 F3-C4 (Figure 12.A), for iPLV and C3-T6 C3-T4 for corrEnv (B).

## 4. Discussion

Our DoCM model identified the dominant coupling modes across experimental time and between every possible pair of EEG sensors as a natural way to reveal the multiplexity of brain oscillations. We revealed a delta synchronisation (increased signal power) and alpha desynchronization (decreased signal power) in SSDs subjects, which was also evidenced by a higher ratio of relative signal power of delta/alpha_1,2_ for SSDs compared to HC. The PD of DoCM untangled trends between the two groups with more pronounced results in amplitude driven dominant coupling modes. Machine learning results revealed an absolute discrimination of SSDs from HC focusing on PD of cross-frequency interactions. This high discriminative power between the two groups is driven by features estimated over amplitude (activity) and alpha frequency driven DoCM (connectivity) which are significant higher in SSDs subjects compared to HC. Focusing on DoCM, we defined a novel nonlinearity index of information flow as the ratio of cross-frequency interactions versus the total number of exist functional interactions, we detected a significantly higher nonlinearity for SSDs compared to HC across the recording time only in the amplitude domain. This result was more pronounced when we focused on alpha frequency. The overall network flexibility was significantly lower in SSD patients compared to HC only in the amplitude domain. Finally, our analysis revealed significant findings for SSDs working with brain activity and the multiplex character of brain connectivity dissociating the distinct role of amplitude and phase coupling modes. Before disentangling one by one the main findings of this study, it is important to mention here that our denoising algorithm based on the combination of ICA and wavelets increased the SNR of EEG recordings. This result supports our findings based on signal power and also on brain connectivity where connectivity estimators are sensitive to SNR levels (see SFigure 1).

Previous studies showed that patients with schizophrenia have reduced delta wave activity also in sleep stages 3 and 4 (Sekimoto et al., 2010) and also during the perception of neutral and emotionally salient words (Alfimova and Uvarova, 2008). Delta synchronisation expressed with an increased EEG delta activity, has been reported in ScZ and bipolar disorders (Howells et al., 2018). Here, we confirmed such a delta synchronisation in SSD patients compared to healthy controls (Figure 3). Increased delta activity (delta synchronization) is associated with central nervous system depression, observed during slow wave sleep, in anaesthesia and in com which are all conditions with characteristic decreased levels of consciousness (Englehardt et al., 1991; Hashemi et al., 2015). A global delta increased activity has been linked to the subthreshold activity of GABAergic neurons originating from the thalamic reticular nucleus and lateral geniculate nucleus (Ulrich et al., 2014; Herrera et al., 2016). Increased delta activity has been linked to lesions in basal forebrain during wakefulness (Fuller et al., 2011) combined with cholinergic and serotonergic blockade (Vanderwolf and Pappas, 1980). Pharmacological models for delta activity have reported interesting findings. The use of an acetylcholinesterase inhibitor in Alzheimer’s disease decreases delta and theta power while it increases alpha power in the left insula following with a cognitive improvement (Gianotti et al., 2008).

In ScZ alpha desynchronization (decreased alpha activity) is reported in adolescent onset ScZ, first-episode ScZ and un-medicated and medicated ScZ (John et al., 2002). We found alpha desynchronization in SSDs subjects compared to healthy controls (Figure 6). It is well-know that desynchronization of alpha activity reflects diverse changes in thalamo-cortical and cortical network communication (Klimesch et al., 1999). Two key subnetworks underlie alpha desynchronization: a) the activation of the visual system, via the reticular activating system (Volavka et al., 1967) as they were revealed by looking at EEG activity recorded during a subject who opens his eyes (eyes-open resting-state) after an eyes-closed resting-state where there is a mass desynchronization of alpha activity and b) desynchronization of alpha activity reflects different changes in thalamo-cortical and cortico-cortical network communication (Klimesch et al., 1999). Decreased alpha activity (alpha desynchronization) has been also associated with central nervous system depression as a consequence of long term use of alcohol, in anaesthesia and vegetative states where all conditions demonstrated decreased levels of consciousness (Hoffman et al., 1995; Kaplan et al., 1985; Lehembre et al.,2012). In healthy conditions, alpha synchronisation is an indicator of healthy resting wakefulness and arousal to attend and also process salient information (Klimesch et al., 1999). Our findings in alpha frequency could be attributed to loss of alertness and arousal with a possible thalamocortical implication. Alpha is generated glutamatergic and muscarinic transmission within the thalamus (Hughes and Crunelli, 2005). Reduced alpha signal power has been seen in cholinergic (Olincy and Freedman, 2012), in glutamatergic (Javitt, 2010) and also in GABAergic (Gonzalez-Burgos and Lewis, 2012) models in schizophrenia.

Studying the ratio of delta/alpha_1,2_, we detected a significantly higher ratio for SSDs compared to HC group similarly to subjects with bipolar disorders and schizophrenia (Figure 7; Howells et al., 2018). Our study is the first one that identified these biomarkers in SSDs subjects. Biophysical models on the source space will further evaluate the origin and the explanation of our observations based on delta and alpha relative signal power.

A consistent decreased functional connectivity pattern in in the α-frequency band has been reported in ScZ. In particular, decreased α connectivity estimated by coherence, lagged coherence and phase synchrony, has been reported at frontal (Di Lorenzo, et al., 2015; Tauscher et al.,1998), fronto-posterior (Di Lorenzo, et al., 2015, Lehmann et al., 2014) and parieto-temporal (Di Lorenzo, et al., 2015) brain areas (for different results see also Andreou et al., 2015, Kam et al., 2013, Winterer et al., 2011, Merrin and Floyd, 1996). Interestingly, two studies reported a high correlation between functional connectivity at rest in the α-frequency band with symptoms of ScZ (Hinkley et al., 2011; Merrin and Floyd, 1996). Contradicting evidence has been reported for fast oscillations in the β-(13– 30 Hz) and γ- (30–200 Hz) frequencies at rest, including both elevated (Di Lorenzo, et al., 2015), reduced (Kam et al., 2013) and intact (Lehmann et al., 2014, Andreou et al., 2015, Tauscher et al., 1998, Winterer et al., 2011) β-band connectivity. Preliminary evidence suggests that β-band functional connectivity is influenced by illness progression and clinical symptomatology (Di Lorenzo, et al., 2015). For a systematic review tailored to functional connectivity evidences using EEG in schizophrenia see Maran et al. (2016).

Here, we adopted our DoCM model to reveal the dominant coupling modes independently in amplitude and phase domain. It is well-known the distinct role of amplitude-to-amplitude coupling and phase-to-amplitude coupling between frequencies (Hyafil et al., 2015). Many previous studies explored the contribution of both low and high frequency oscillations to explain a range of cognitive deficits in schizophrenia modulated with specific frequency content (Moran and Hong,2011). Here, we analysed phase-to-phase (within frequencies) with phase-to-amplitude couplings (cross-frequencies) in the phase domain and amplitude-to-amplitude couplings of the envelopes of either the same frequency (within frequencies) or of different frequencies (cross-frequencies). Phase based intrinsic coupling modes are band-limited in specific frequency bands, are extended from local to large-scale coupling networks and changed in disorders with structural or even functional network alterations (Engel et al., 2013). Envelopes of Amplitude based intrinsic coupling modes display a typical frequency range on the ultra-slow spectrum below 0.15 Hz similar to BOLD activity. We typically estimated correlation of the envelope between frequency-dependent brain signals but the spectrum of the envelopes is within that frequency scale. Amplitude based intrinsic coupling modes are extended both locally and globally (network level) and might be severely affected in disorders with a predominant structural network alteration (Engel et al., 2013). Structural brain changes in subtypes in schizophrenia could further support our findings in amplitude driven DoCM which are more sensitive in structural changes (Zhang et al., 2014).

Comparing the findings of PD of DoCM with phase driven (Figure 8) and amplitude driven (Figure 9) frequency-interactions modes, we untangled a reconfiguration of DoCM in the latter case in SSDs subjects driven by alpha sub-bands. Attention can be allocated into a dorsal, top-down network and a ventral, bottom-up network. The top-down network is responsible for attention to certain features while the bottom-up network is mainly stimulus-driven. These networks are identified in both task-based (Corbetta and Shulman, 2002) and resting state neuroimaging paradigms (Fox et al., 2006) and are directly related to attentional functions that are active even at resting-state conditions. Both types of cross-frequency phase–amplitude coupling (PAC) and amplitude–amplitude coupling (AAC) are highly preserved in top-down and bottom-up networks in macaque auditory cortex (Marton et al., 2019). Clearly, the most discriminative features of PD of DoCM supported an absolute accuracy the two groups. Selected features revealed also a complementary role of theta modulating DoCM with higher frequency bands (beta/gamma), an observation that has been reported also in a resting-state study in HC with MEG modality (Florin and Baillet, 2015). Additionally, we observed a significant lower global FI in SSDs compared to HC only in amplitude driven DoCM. In a recent lifespan study with healthy controls, we found aberrant FI for two groups, a dyslexic group and a mild traumatic brain injury (Dimitriadis et al., 2019). FI can be seen as a global index of the normal multiplexity behaviour of nested oscillations. DoCM, their PD and other features, where some of them are presented here, are further needed to explain in more detail group differences between healthy controls and targeted groups.

The Nonlinearity Index defined and reported here for the first time showed a higher nonlinearity of resting-state networks for SSDs subjects compared to HC only in amplitude domain while in phase there was no group difference. Our findings have been consistent both using the whole repertoire of within and between frequencies couplings but also focusing on alpha sub-bands modulators (Figure 11). Our study first dissociate the potential different role of amplitude and phase driven intrinsic coupling modes in general and specifically in the target group of SSDs. These results are support by findings of a recent study that underlined the complementarity of cortical phase and amplitude coupling patterns. This study revealed also the importance of amplitude coupling measures (Siems and Siegel, 2020). Our results untangled an interesting role of alpha frequency as the key frequency modulator that demonstrates a similar behavioural role in phase coupling modes in both groups and a distinct role in amplitude domain dissociating its role in SSDs from healthy controls. Further research is needed based on biologically inspired models like dynamic causal modelling that will attempt to explain this modulator role of alpha amplitude in SSDs under the spectrum of GABAergic inhibition and disinhibition that it is well know that modulate cortical synchrony in low brain frequencies (Xiao et al., 2012; Shaw et al., 2019). GABA neurotransmission is altered in schizophrenia playing a key role (Schmidt and Mirnics,2015). Overall, this finding can be seen as a continuous and intense alertness of attentional systems in SSDs subjects (Klimesch et al., 1999). Our analysis focused on the eyes-closed task, a condition that produced more uniform and consistent findings in psychiatric disorders (Newson and Thiagarajan, 2019) than eyes-open resting-state.

We found group differences in inter-hemispheric links between the following sensors pairs:O1-P4 P3-O2 P3-T6 C3-T6 F3-O2 F3-C4 for iPLV and C3-T6 C3-T4 for corrEnv. Group-averaged iPLV values were higher for SSDs while group-averaged corrEnv were higher for healthy controls. Our findings are in conjunction with a previous study using EEG resting-state in schizophrenia (Olejarczyk and Jernajczyk2017).

This study is the first one that explored simultaneously both intra-frequency and crossfrequency interactions in both amplitude and phase domain in SSDs under DoCM model. Under this framework, we can decipher the multiplexity of complex electrophysiological aberrant connectivity observed in SSDs by integrating the available intrinsic coupling modes.

Our DoCM model untangled the multiplexity of human brain dynamics recorded with EEG by integrated both within and between frequencies coupling modes under the same framework (Buzsaki and Watson, 2012). DoCM model hypothesizes that the fluctuation of DoCM can capture the complexity of functional brain connectivity during both spontaneous and cognitive tasks. The transitive nature of DoCM across expertimental time can be quantified with FI which is a direct mearure of brain’s multiplexity. FI can be seen as a reflex index that describes the readiness of the brain to respond to new stimuli.

It would be interesting to follow the same methodology during cognitive tasks and also on the source level in order to get the advantage of animal models, pharmaco-based studies and fMRI studies that revealed the mechanistic explanation of aberrant networks in schizophrenia (Hunt et al., 2017).

The whole study is unique, innovative and pioneering in terms of analytic pathway and scientific results. However, there are two basic limitations. The first refers to the interpretation of the results on the EEG surface level instead of virtual cortical sources. It would be interesting to follow the same methodological approach in an EEG study recorded healthy controls and SSDs using a large number of EEG net sensors that support the source reconstruction approach. The second drawback is the limitation of the free EEG database that involves only recordings without any access to neuropsychological assessment battery. This missing part prevented us to correlate the novel chronnectomic and semantic features with trivial neuropsychological estimates.

## 5. Conclusions

In the present study, we adopted a holistic approach of exploring how brain activity and connectivity mediated by intra and cross-frequency interactions differentiate in SSDs subjects compared to HC. We untangled a reconfiguration of amplitude driven DoCM in SSDs subjects mediated by alpha activity. Our findings detected significant and novel findings that will help clinicians to detect SSDs with a low cost EEG device while they can design proper neurofeedback trainings tailored to DoCM instead of only relative signal power.

## Data Availability

The study analyzed an existing free EEG database that can be downloaded from the following link:
http://brain.bio.msu.ru/eeg_schizophrenia.htm

## Abbreviations

PLEs: psychotic-like experiences
SSDs: Schizophrenia Spectrum Disorders
DFCG: dynamic functional connectivity graph
iPLV: imaginary part of phase lag value
corrEnv: correlation of the envelope
iDFCG: integrated dynamic functional connectivity graph
DoCM: dominant coupling mode model
PD: probability distribution
FI: Flexibility Index
RSP: relative power spectrum
CFC: cross-frequency coupling

## Acknowledgments

I would like to thank Professor David Linden for his careful proof-reading, his thoughtful comments based on his expertise that finally improved and shaped the final paper.

I would like also to thank Professors Kaplan AI, Gorbachevskaia N L & Kozlova I. for making this dataset available to the public for further research.

SID was supported by MRC grant MR/K004360/1 (Behavioral and Neurophysiological Effects of Schizophrenia Risk Genes: A Multi-locus, Pathway Based Approach). SID is also supported by a MARIE-CURIE COFUND EU-UK Research Fellowship. We would like to acknowledge RCUK of Cardiff University and Wellcome Trust for covering the publication fee.

## Conflict of Interest

The authors declare that the research was conducted in the absence of any commercial or financial relationships that could be construed as a potential conflict of interest.

## Notes

### Competing Interest Statement

The authors have declared no competing interest.

### Clinical Trial

The study has been approved within the Hospital of Moscow and not in a global repository.
The study analyzed a free available dataset that can be downloaded from here:
http://brain.bio.msu.ru/eeg_schizophrenia.htm.

